# A link between inflammatory biomarkers and lung ultrasound observations in patients with SARS-CoV-2 infection

**DOI:** 10.1101/2020.06.22.20134270

**Authors:** Aurelio Luis Wangüemert Pérez, Juan Marco Figueira Gonçalves, José María Hernández Pérez, Yolanda Ramallo- Fariña, José Carlos del Castillo Rodríguez

**Author notes:** **Correspondence:** Aurelio Luis Wangüemert Pérez, San Juan de Dios Hospital, Ctra. Santa Cruz - Laguna 53, 38009, Santa Cruz de Tenerife, Spain.

## Abstract

Lung ultrasound (LUS) has shown to correlate well with the findings obtained by chest computed tomography (CT) in acute-phase COVID-19. Although there is a significant correlation between blood biomarkers and CT radiological findings, a potential correlation between biochemical parameters and LUS images is still unknown. Our purpose was to evaluate a potential association between lung lesions visualised by LUS and blood biomarkers as well as the ability to predict mortality from two different lung ultrasound scoring systems (LUSS). We performed a retrospective observational study on 45 patients aged >70 years with SARS-CoV-2 infection who required hospitalisation. LUS was carried out at admission and on day 7, when the clinical course was favourable or earlier in case of worsening. Disease severity was scored by means of LUSS in 8 (LUSS8) and in 12 (LUSS12) quadrants. LUS and blood draw for inflammatory marker analysis were performed at the same time. The correlation between biochemical parameters and either LUSS score was significant for ferritin levels. It was 0.486 (p=0.001) for LUSS8 and 0.458 (p=0.002) for LUSS12. Using a threshold score of 15 with LUSS12 predicted mortality in 86.7% of cases (OR_crude_ 31, CI 95% 4.79–200.51). Applying a threshold of 10 with LUSS8 predicted mortality in 88.9% (OR_crude_ 69.75, CI 95% 6.90–705.20). There is a correlation between ferritin levels and LUSS. The prognostic capacity of LUSS12 does not surpass that of LUSS8.

**“Take-home message”:** There is a correlation between lung ultrasound scoring results and serum ferritin levels in patients with SARS-CoV-2 infection.

## Introduction

The World Health Organization (WHO) has declared COVID-19 a pandemic [1]. Although current evidence suggests that most infections manifest mildly, up to 16% of cases may require hospital admission for developing severe pneumonia with acute respiratory distress syndrome (ARDS), sepsis and even septic shock [2]. A large study carried out in Spain with more than 12 000 patients showed a mortality rate of less than 2% in patients under 40 years of age, but values from 25% to 50% in individuals over the age of 70 years, which highlights the vulnerability of the elderly [3].

Due to its high sensitivity and specificity, lung ultrasound (LUS) is considered a suitable alternative to conventional radiological methods to diagnose pneumonia. Various scientific societies have highlighted its utility in acute-phase COVID-19, as the resulting data showed a good correlation with those from high-resolution computed tomography (HRCT) of the chest. Based on these findings, the technique has gained importance both for disease diagnosis as well as follow-up, as it can facilitate daily monitoring with no need for radiation or intra-hospital patient transfer [4–8].

Given the unpredictable evolution of the disease, multiple research has evaluated indicators that may correlate with a more unfavourable course. Radiological patterns and their extent in COVID-19 have been described as being of prognostic value [9,10]. In addition, different serum biomarkers, such as leukocyte and lymphocyte counts, lactate dehydrogenase (LDH) levels, D-dimer, procalcitonin, troponin I and ferritin seem to provide information on the severity of the process [11–16]. Although scientific evidence correlates serum biomarkers with chest computed tomography (CT) findings [17], there are no studies yet to evaluate any correlation between biochemical parameters and LUS imaging.

The aim of our study was to evaluate potential correlations of the extension and severity of LUS-visualised, SARS-Cov-2 induced lung lesions and serum biomarkers in patients over 70 years of age, on the one hand, and the mortality prediction capacity of two lung ultrasound scoring systems (LUSS), on the other hand.

## Methods

### Study design and participants

This was an observational study with retrospective follow-up of patients older than 70 years with SARS-CoV-2 infection, who were admitted to a secondary hospital in the island of Tenerife (Spain) in the period of 1 February 2020 to 15 May 2020. Diagnosis was made in all patients by means of real-time reverse-transcription PCR of SARS-CoV-2 RNA from nasopharyngeal smears. The only exclusion criteria was when LUS and blood draw did not coincide on the same day.

### Variables

Patient demographic and comorbidity data were collected at hospital admission. All patients underwent two ultrasound examinations, the first on admission and the second on day 7 in case of a favourable clinical course, but the interval was shortened in the event of clinical deterioration. Extent and severity of the ultrasound findings was determined by using two scoring protocols, LUSS with 8 quadrants (LUSS8) and LUSS with 12 quadrants (LUSS12) [18,19]. Both protocols assign scores of 0–3, where 0 stands for pattern A, 1 for pattern B (i.e. non-confluent B-lines), 2 for confluent B-lines and 3 for dense and largely extended white lung with or without consolidation (pattern C) [20].

In the LUSS8 protocol, 8 locations were analysed. They consisted of 4 quadrants in each hemi-thorax (the anterior region of the chest with its upper anterior and lower anterior quadrant and the axillary thorax with its upper lateral and lower lateral quadrants). Scores ranged from 0 to 24.

In the LUSS12 protocol, 12 quadrants were analysed. The upper and lower quadrants of the posterior thorax were added to the locations mentioned above, and 6 zones were explored in each hemi-thorax. Scores ranged from 0 to 36.

Ultrasound scans and scoring were performed by a single operator experienced in LUS. A Philips EnVisor C ultrasound scanner (Philips Eindhoven, The Netherlands), equipped with a 3–5 Mhz convex probe or a 5–13 Mhz linear probe, was used according to the patient’s physical characteristics and the region to explore. Images and videos were recorded on a computer.

Together with ultrasound evaluation, blood samples were obtained to determine leukocyte and lymphocyte numbers, D-dimers, ferritin, procalcitonin and C-reactive protein (CRP).

### Ethics

The study was approved by the Research Ethics Committee of the University Hospital Nuestra Señora de la Candelaria.

### Statistical analyses

Continuous variables were expressed as means±SD. To compare means of two groups of independent samples, Student’s t-test was applied. OR_crude_ (crude odds ratio) was obtained by univariate logistic regression between the variable “mortality” and the dichotomised scores according to their cut-off point. ORs (odds ratios) were accompanied by their 95% CI (confidence interval). Following Manivel et al. [21], a cut-off point of ≤15, 15> was applied for LUSS12. For LUSS8, the cut-off point was obtained taking into account the distribution of data in relation to mortality. Statistical significance was set at 5% and a trend towards significance accepted for p<0.1. Analyses were conducted with SPSS software (version 21; SPSS, Chicago, IL, USA).

## Results

### Study population

A total of 45 patients were included. Their baseline characteristics are shown in Table 1. Women accounted for the majority (55.6%) of patients. The mean age was 82±9.9 years, and the most frequent comorbidities were arterial hypertension (66.7%), dyslipidemia (62.2%) and Type 2 diabetes mellitus (42.2%). A total of 10 patients (22%) died during hospital stay. The deceased had a higher prevalence of hypertension, dyslipidemia and ischaemic heart disease than the survivors (p<0.05). The mean follow-up of the survivors was 20.2±4.7 days and of the group of deceased patients 7.4 ± 5.4 days.

**TABLE 1.** Baseline characteristics of all included patients

|  |  |
| --- | --- |
| Patients n | 45 |
| Male n (%) | 20 (44.4) |
| Age years (mean±SD) | 82.4±9.9 |
| Arterial hypertension n (%) | 30 (66.7) |
| Type 2 diabetes mellitus n (%) | 19 (42.2) |
| Chronic obstructive pulmonary disease n (%) | 12 (26.7) |
| Dyslipidemia n (%) | 28 (62.2) |
| Chronic kidney disease n (%) | 11 (24.4) |
| Moderate-severe cognitive decline n (%) | 7 (15.9) |
| Hypothyroidism n (%) | 6 (13.3) |
| Chronic atrial fibrillation n (%) | 2 (4.4) |
| Neoplasms (solid tumour, leukaemia, lymphoma) n (%) | 2 (4.4) |
| SD: standard deviation. |  |

### Correlation between biochemical parameters and LUSS scores

The correlation between the analysed biochemical parameters and LUSS8 and LUSS12 scores was only significant for ferritin levels and was 0.486 (*p*=0.001) and 0.458 (*p*=0.002), respectively. A trend towards significance was detected for CRP with a correlation of 0.269 (*p*=0.074) for both scoring systems (table 2).

**TABLE 2.**
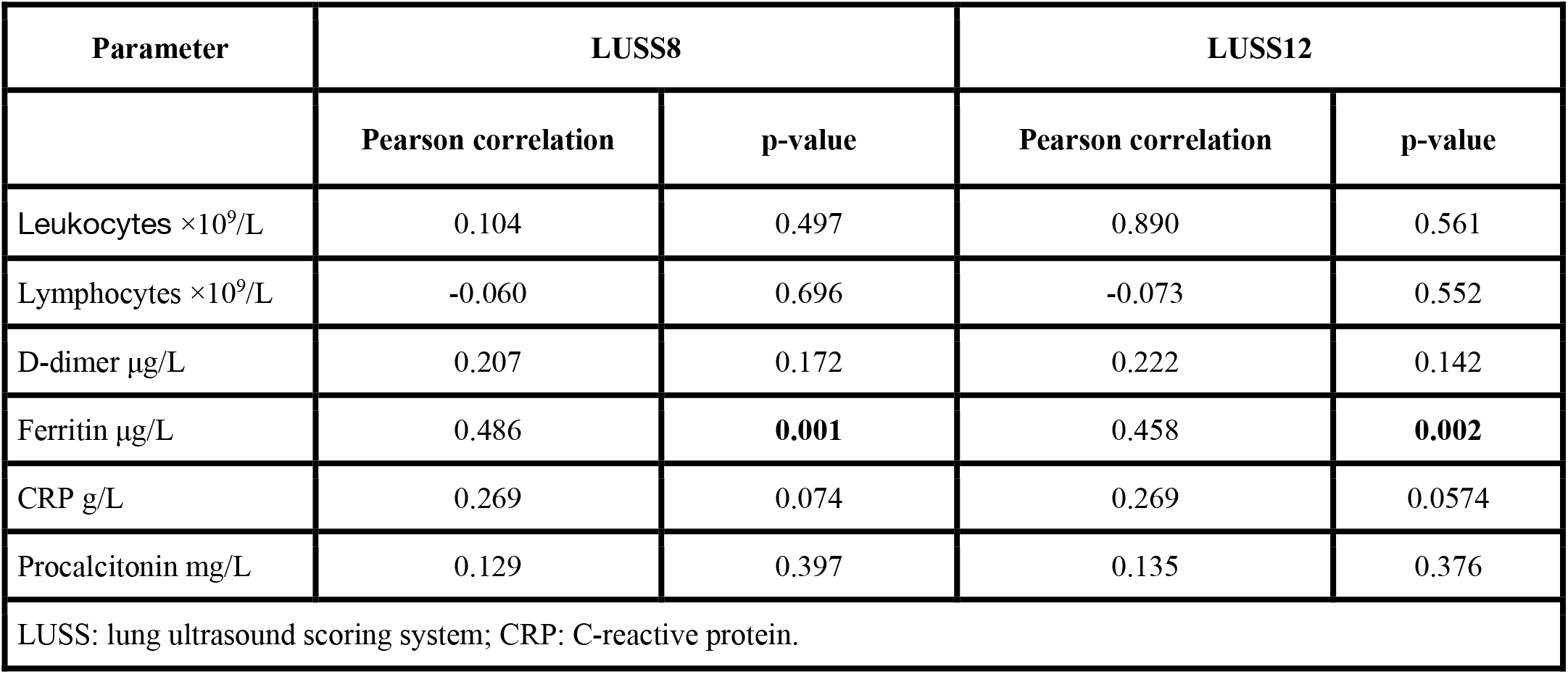
Correlation between blood parameters and lung ultrasound scoring systems

Taking into account the cut-off scores, the difference in means of the biochemical parameters was only significant for ferritin. In patients with LUSS8 scores >10, ferritin levels were higher than in patients with scores ≤10, i.e. 664.6±445.0 μg/L and 245.13±207.6 μg/L (p=0.008), respectively. As to LUSS12, ferritin levels were 254.9±204.2 μg/L for patients with scores ≤15 and 700.4±448.3 μg/L for those with a scores >15 (table 3).

**TABLE 3.** Relationship between lung ultrasound scoring system cut-off points, blood parameters and mortality

| LUSS12 |  |  |  |  |
| --- | --- | --- | --- | --- |
| Blood parameter | LUSS12 ≤15<br>n=33 | LUSS12 >15<br>n=12 | t-Student's | p-value |
|  | Mean (SD) | Mean (SD) |  |  |
| Leukocytes ×10<sup>9</sup>/L | 6.79 (3.26) | 8.20 (3.91) | -1.22 | 0.229 |
| Lymphocytes ×10<sup>9</sup>/L | 138.39 (786.58) | 1.19 (0.59) | 0.6 | 0.552 |
| D-dimer µg/L | 1 157.79 (2 643.68) | 1 492.25 (1 905.53) | -0.401 | 0.691 |
| Ferritin µg/L | 245.94 (204.24) | 700.36 (448.27) | -3.248 | **0.007** |
| CRP g/L | 3.58 (5.24) | 8.94 (8.39) | -2.073 | 0.057 |
| Procalcitonin mg/L | 1.1 (0.4) | 1.4 (1) | -1.07 | 0.305 |
| LUSS 8 |  |  |  |  |
| Blood parameter | LUSS8 ≤10<br>n=32 | LUSS8 >10<br>n=13 | t-Student's | p-value |
|  | Mean (SD) | Mean (SD) |  |  |
| Leukocytes ×10<sup>9</sup>/L | 6.83 (3.30) | 7.99 (3.82) | 1.018 | 0.314 |
| Lymphocytes ×10<sup>9</sup>/L | 142.68 (798.77) | 1.17 (0.57) | -0.634 | 0.529 |
| D-dimer µg/L | 1 183.28 (2 681.86) | 1 403.77 (1 852.09) | 0.27 | 0.788 |
| Ferritin µg/L | 245.13 (207.57) | 664.58 (445.02) | 3.136 | **0.008** |
| CRP g/L | 3.57 (5.32) | 8.55 (8.16) | 2.035 | 0.058 |
| Procalcitonin mg/L | 1.1 (0.4) | 1.4 (1) | 1.023 | 0.324 |
| Mortality |  |  |  |  |
|  | Survivors<br>n=35 | Deceased<br>n=10 | t-Student's | p-value |
|  | Mean (SD) | Mean (SD) |  |  |
| Score from LUSS12 | 6.89 (5.91) | 19.30 (4.32) | 6.171 | **<0.0001** |
| Score from LUSS8 | 4.11 (3.93) | 13.0 (3.59) | 6.416 | **<0.0001** |
| Variation LUSS12* | -0.69 (2.7) | 3.90 (3.48) | -4.43 | **<0.0001** |
| Variation LUSS8* | -0.11 (2.23) | 3.20 (3.22) | -3.05 | **0.011** |
| LUSS: lung ultrasound scoring system; CRP: C-reactive protein; SD: standard deviation; *difference between scores from the first and second ultrasound scan. |  |  |  |  |

### Association between mortality and LUSS scores

As compared to survivors, patients who died had higher scores both in LUSS8 (13±3.59, p<0.0001) as well as LUSS12 (19.3±4.32, p<0.0001). When applying LUSS12, a cut-off score of 15 correctly predicted mortality in 86.7% of cases (OR_crude_ 31.0, 95% CI 4.79–200.51). With regard to LUSS8, a cut-off score of 10 accurately identified mortality in 88.9% of the patients (OR_crude_ 69.75, 95% CI 6.90–705.20). Furthermore, an increase in scores from both systems during follow-up was associated with higher mortality with p=0.01 and p<0.0001, respectively (table 3).

## Discussion

We draw the following two conclusions from this study: First, there is a correlation between serum parameters (ferritin) and the degree of lesions in the lungs as identified by LUS in patients over 70 years of age with SARS-CoV-2 infection. Second, the ability to predict mortality from a 12-quadrant LUSS does not surpass 8-quadrant scanning.

In acute-phase COVID-19, a good correlation between LUS and thoracic HRCT data has been reported, which emphasises the importance of LUS in both diagnosis and follow-up [7,8]. Laboratory parameters, such as leukocyte and lymphocyte numbers, LDH, D-dimer, procalcitonin, troponin I and ferritin levels are related to the clinical course of the infection and thus can provide information about a need for intensive care unit admission or the risk of mortality [11–16]. Various studies have shown a worse course of COVID-19 in patients with higher levels of interleukin-6 and ferritin than in individuals with a milder disease course [14–15], so that both biomarkers were suggested for patient monitoring during their hospital stay [16]. A rise in these parameters is related to a systemic inflammatory response syndrome (SIRS) [22], a so-called cytokine storm, which can progress into ARDS, darkening these patients’ prognosis. Chen et al. [13] detected higher levels of CRP, ferritin, LDH and D-dimer in severe cases of COVID-19 compared to a milder ones. On CT scans, patients with COVID-19 generally present chest opacities, the extent of which has been correlated with laboratory parameters. A study published by Xiong et al. [17] found that elevated CRP and LDH levels correlated with the extent of pneumonia quantified by chest CT.

In our study, elevated ferritin levels correlated with high LUSS scores, which may be an expression of lung damage in the context of SIRS. Such exaggerated inflammatory response was shown to be a prognostic factor in COVID-19 patients [12]. In this sense, ultrasound findings from lung parenchyma scans are plausible to reflect disease severity and may therefore offer information on the course of the process from the very beginning.

On the other hand, we would like to underline that the mortality prediction capacity was similar when and independent of exploring either 8 or 12 quadrants. This data indicates that disease progression could be evaluated by only exploring the anterior and lateral quadrants. The aspect acquires special relevance in patients where, due to their clinical situation, the posterior thoracic field cannot be explored. Furthermore, the need to explore fewer quadrants reduces the exposure time of the medical staff and therefore the risk of contagion.

To our knowledge, this is the first study to evaluate the relationship between blood biomarkers and the extent of pleuro-pulmonary ultrasound findings in patients hospitalised for SARS-CoV-2 infection. We must put emphasis on the age range of our patients, as—although they form the most vulnerable group with a high mortality rate—only few studies have focused on this group yet [23–24]. One of the limitations of this work is that a larger sample size could potentially have revealed correlations with other biomarkers. Furthermore, the examination was performed by a single operator, so that we cannot rule out inter-observer differences when sonographic scoring is performed by more than one person.

In conclusion, there is a correlation between ferritin levels and the scores obtained by LUSS in hospitalised patients older than 70 years with SARS-CoV-2 infection. In this study, high scores correlated with high levels of the biomarker. In addition, the prognostic capacity of scanning 12 fields does not seem advantageous over 8 quadrants, which allows reducing the staff exposure time and thus the risk of contagion. Studies with a larger sample size are necessary to confirm these findings.

## Data Availability

There is availability of all the data to which it refers

